# Inter-step Variations of Stairways and Associations of High-Contrast Striping and Fall-related Event Observations

**DOI:** 10.1101/2023.01.28.23285143

**Authors:** Chayston B. Brown, Shandon L. Poulsen, Tyson S. Barrett, Christopher J. Dakin, Sara A. Harper

**Author notes:** Correspondence: Sara A. Harper, Address: 7000 Old Main Hill, Utah State University, Logan, UT, 84322, USA, Twitter: @sharpe18.

## Abstract

**Background:** Variability in the riser and depth of each step in a stairway can decrease step predictability, and increase users’ fall risk. Unfortunately, inconsistencies in inter-step riser height and depth are common, but their impact may be lessened by adding high-contrast tread edges and vertical, monochrome striping applied to the bottom and top steps’ faces. Such contrast enhancement may draw greater attention to the steps’ edges or enhance the precision of the edge’s estimated location.

**Purpose:** To determine if greater inconsistency in inter-step riser height and depth are associated with more frequent slips, trips, and falls, and if these events are reduced in flights of stairs with contrast-enhanced step edges.

**Methods:** Stair users were videotaped on two public stairways. One stairway had black vinyl strips applied to the steps’ edges and black-and-white vertical stripes on the first and last steps’ faces. The intervention stairway was switched halfway through data collection. Stair users were coded for whether they experienced a fall-related event. A Monte Carlo simulation was used to determine the probability of observing a range of plausible distributions of fall-related events.

**Results:** Flights of stairs with riser height variability of 14 mm and tread depth variability of 38 mm were associated with 80% of the observed fall-related events. 13 of 16 (81%) fall-related events occurred on the control stairway compared to 3 of 16 (19%) on the intervention stairway. The distribution of fall-related events observed between conditions had a probability of occurring by random chance of less than 0.04.

**Conclusion:** These data suggest a vision-based strategy (i.e., striping) may counteract fall risk associated with inter-step riser height and depth inconsistencies. While the mechanisms of its action remain unclear, the high-contrast striping appears to reduce the incidence of fall-related events in the presence of inter-step riser height and depth inconsistencies.

## 1. Introduction

When approaching a stairway, stair users seemingly anticipate uniformity in the riser height and tread depth of steps (Archea et al., 1979; Nemire et al., 2016). However, as a result, the reduced attentional resources allocated to estimating these metrics could potentially hinder safe negotiation of the stairway. Unfortunately, inter-step height and tread depth inconsistencies are common, and inconsistencies of just 6.35 mm can cause a disruption in step cadence, and increase fall risk or the likelihood of an accident (Templer, 1992; Wright & Roys, 2005).

Here we sought to evaluate whether an intervention, namely the addition of black stripes to the top, front edge of each step, could counteract the impact of inter-step inconsistencies on the number of observed slips, trips, and falls (fall-related events). As described previously (Harper, 2022), Elliott, Foster, and colleagues have demonstrated that vertical, monochrome striping on the face of the bottom and top steps can increase vertical foot clearance (Elliott et al., 2015; Elliott et al., 2009; Foster et al., 2016; Foster et al., 2015a; Skervin et al., 2021). Adding high-contrast visual strips along the top front edge of all the steps (Cohen & Sloan, 2016; den Brinker et al., 2005; Elliott et al., 2015; Foster et al., 2014a; Thomas et al., 2021; Zietz et al., 2011) could lead to increases in heel clearance.

We hypothesized that flights of stairs with greater inter-step inconsistency in riser heights and tread depths would have a greater number of fall-related events compared to stairs with fewer inter-step riser heights and tread depths. Also, that the addition of high-contrast striping to the step’s top front edge and vertical, monochrome striping to the face of the first and last would reduce fall risk on stairs with greater inter-step inconsistencies compared to stairs with similar variability but without the intervention.

To test our hypotheses, we estimated the probability of observing a range of distributions of fall-related events that could plausibly occur by chance given our hypotheses [high/low riser height and tread depth variations, and control/striping intervention conditions] (see **Figure 1**).

**Figure 1.**
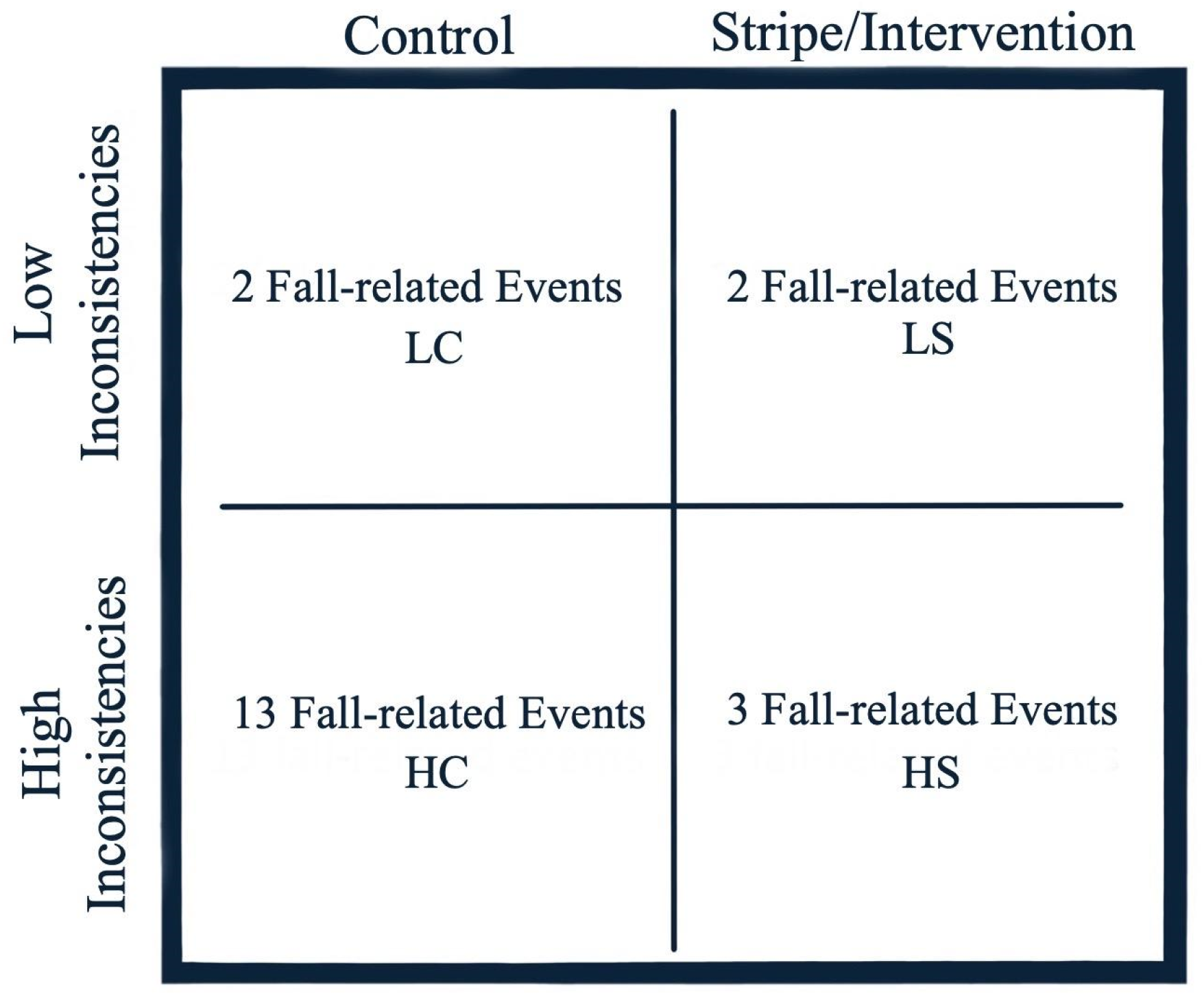
The following assumptions evaluated the observed fall-related event distribution. Abbreviations: high variations, control condition (HC); high variations stripe intervention (HS); low variations, stripe intervention (LS); low variations, control condition (LC). Four assumptions were used to code our hypotheses based on our a priori knowledge of the total number of fall-related events recorded (20): (1) the number of fall-related events HC > LC. (2) the difference between LS and LC < 2 fall-related events. (3) the number of falls in HC will be ≥ two times of HS. (4) the difference between LC and HC > the difference between LS and HS. The probability of a distribution meeting these assumptions occurring by chance is approximately 0.0358 with a sample size of 20.

We codified our hypotheses in the Monte Carlo simulation using four constraints based on our hypotheses as follows.

1. There would be more fall-related events on control condition (without the contrast intervention) flights of stairs with greater variability than those with lower variability.
2. If falls were largely due to greater inter-step inconsistencies, then both control and intervention (with the contrast enhancement) flights of stairs with low variability should observe a comparable frequency of fall related events (i.e., less than two fall-related events difference between conditions).
3. If the intervention acts to reduce the frequency of observed falls then we should observe fewer (less than or equal to half) falls on the high variability intervention stairs compared to the high variability control stairs.
4. If the intervention acts to reduce the frequency of falls then the relative difference in fall-related events between high and low-variability stair flights in the control condition should be greater than that of the intervention flights, due to the effect of the intervention.

## 2. Material and methods

### 2.1 Participants

This study took place on two public stairways at Utah State University. Consequently, most stair users appeared to be college-aged adults.

### 2.2 Protocol

High resolution security cameras (8 megapixels, 4K Ultra HD, 3840×2160 resolution at 7 frames per s [Lorex cameras, Lorex Technology Inc., Markham, CA, USA]) were placed in the stairways to record stair users’ behavior.

### 2.3 Intervention

High-contrast black vinyl film (Gerber High Performance Series 220 vinyl film, Gerber Technology, Tolland, CT, USA) stripes (5.5 cm wide, 0.063-0.09 mm thickness) were applied flush to the top front edge of each stair (Elliott et al., 2015; Foster et al., 2016; Foster et al., 2014b). 19 black-and-white vertical vinyl stripes (12 cycles/1 meter) were placed on the very bottom and top steps’ faces (Elliott et al., 2015; Foster et al., 2016; Foster et al., 2015b). The stairways had inconsistent inter-step height and width variation, ranging from: 5 mm of variation in the East, upper flight, to 14 mm in the East, lower flight, to 12 mm in the West, upper flight, and finally 14 mm in the West, lower flight. There was also inter-step width variation ranging from: 4 mm in the East, upper flight, to 38 mm in the East, lower flight, to 6 mm in the West, upper flight, and 38 mm in the West, lower flight (see **Figure 2**). The control stairway was unaltered and used to compare to the intervention stairway. Halfway through data collection, the intervention (striped) and control stairways were switched (i.e., counterbalanced).

**Figure 2.**
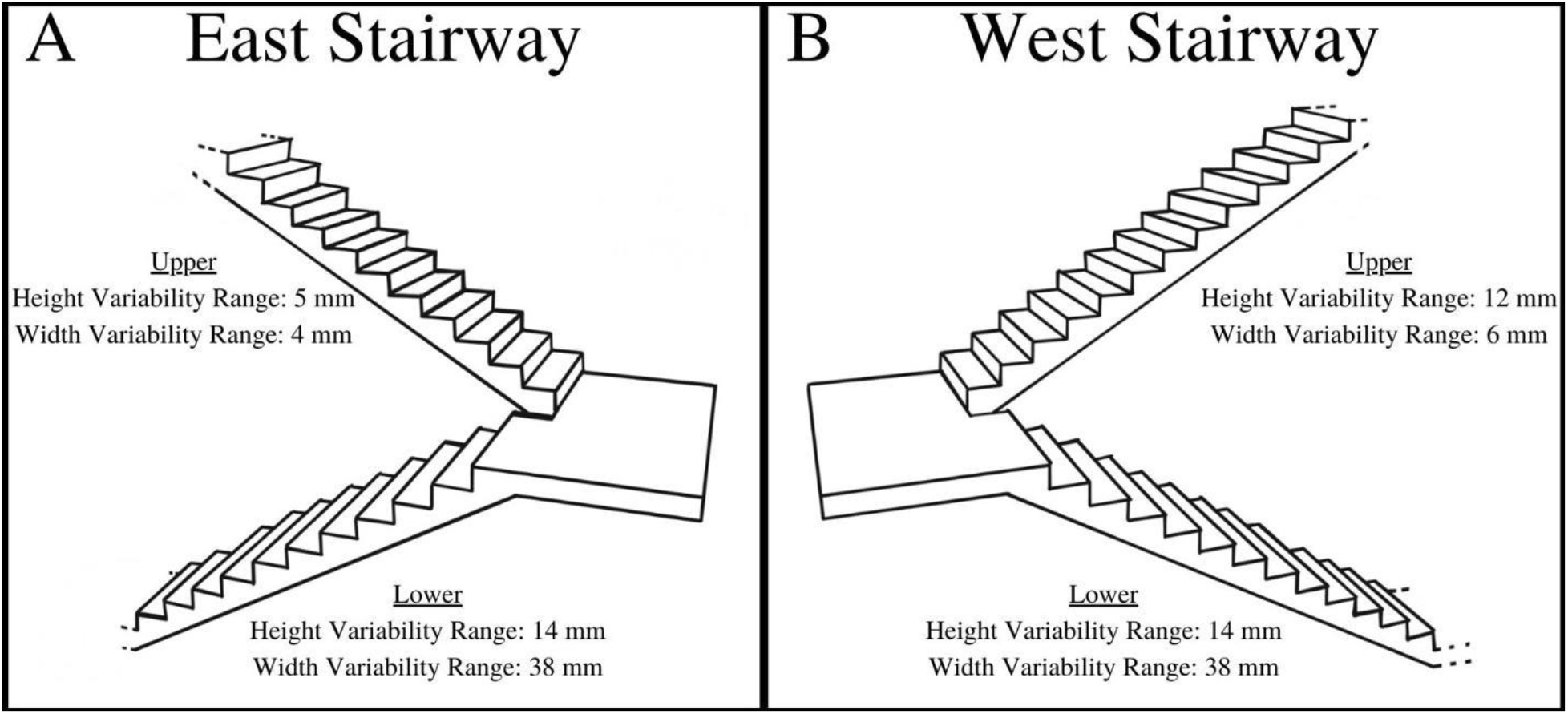
East upper and lower stairway flights inter-step riser height inconsistencies ranged of 5 mm and 14 mm, and inter-step depth of 4 mm and 38 mm respectively (A). West upper and lower stairway flights inter-step riser height inconsistencies ranged of 12 mm and 14 mm, as well as inter-step depth of 6 mm and 38 mm respectively, are shown (B).

### 2.4 Measures

Each outcome variable was assessed by stairway location (East, West) and condition (intervention, control), as well as by stairway flight (lower, upper). We coded stair users’ navigation direction (ascent, descent), and the presence of a fall-related event. If a fall-related event occurred, stairway and step number (starting from the bottom to the top) were recorded. Data were encoded using Microsoft Excel (Microsoft Corporation, Redmond, WA, USA).

### 2.5 Statistical Analysis

Data are presented as mean ± standard deviation as well as count (percentage) of observed results. The distribution of fall-related events was assessed across stairway flights (inter-step inconsistencies), and condition (control, intervention) using a ½ inch threshold (∼13 mm) given that 75% stair accidents occurred in stairways with step riser height inconsistencies of ≥ 12.7 mm (Miller, 1958). A Monte Carlo simulation in Julia (Bezanson et al., 2017) was used to estimate the probability of observing the distribution of fall-related events defined by our hypotheses by chance.

## 3. Results

While the average step riser height for all steps of the East and West stairways measured at the center of the steps were 168 ± 4 mm, the average riser height of the West stairway steps was 170.9 ± 3.4 mm, and those of the East stairway was 166.0 ± 3.0 mm, independently. The average step depth for the East and West stairways was 327.93 ± 7.7 mm, the average depth of the West stairway steps was 327.4 ± 7.5 mm, and the Easy stairway steps was 328.6 ± 8.1 mm, respectively.

Of twenty observed fall-related events, 16 of 20 (80%) events were observed on the flights where inter-step inconsistencies were the greatest (riser height range 14 mm, depth range 38 mm). Between East and West stairways, seven of twenty (35%) fall-related events occurred on steps that had a step riser height greater than one standard deviation from their mean including: two fall-related events that occurred on the first step while ascending (step riser height: 170 mm, flight mean and standard deviation: 166.0 ± 3.0 mm), three events on the last step while descending the East, lower stairway (155 mm, 166.0 ± 3.0 mm), one event on the last step while ascending the East, upper stairway (170 mm, 166.0 ± 3.0 mm), and one event on the second-to-last step while ascending the West, lower stairway (173 mm, 170.9 ± 3.4 mm).

On flights where step riser height ranged 14 mm and step depth ranged 38 mm, 13 of 16 (81%) fall-related events occurred on the control stairway condition (no striping), compared to 3 of 16 (19%) on the striped intervention stairway. The estimated probability of observing data that fit the range of distributions constrained by our hypotheses, and using sample size of 20, was approximately 0.0358 (see **Figure 1**). This result suggests that a) inter-step inconsistencies may be contributing to falls, and b) that adding a striping intervention to the stairs may reduce the impact of inter-step inconsistencies on fall-related events.

## 4. Discussion

We sought to assess the impact of inter-step riser height and tread depth inconsistencies on fall-related events (e.g., slips, trips, or falls) on stairways, and to determine whether fall risk may increase with greater inter-step inconsistencies on unaltered stairways (i.e., control condition) compared to a striping intervention. The lower flights of stairs, that had inter-step riser heights that ranged 14 mm and inter-step depths that ranged 38 mm, accounted for 16/20 (80%) of the observed fall-related events, supporting the notion that stairways with greater inter-step inconsistencies may be associated with a greater risk for fall-related events. Furthermore, 7/20 (35%) fall-related events were observed on steps where step riser height was greater than one standard deviation from the flight mean. This evidence supports the hypothesis that flights of stairs with greater inter-step height inconsistencies would exhibit a greater number of fall-related events compared to flights with lower inter-step inconsistencies. And, that the addition of high-contrast striping to the stairs may reduce the impact of greater inter-step inconsistencies.

Even small variations in riser height and tread depth can have a negative impact on stairway falls. Variations in step riser height as small as 6.35 mm can cause a disruption in step cadence (Templer, 1992), and thus increase fall risk. Moreover, a review of 80 stairway falls from 1992 to 2007 found that 60% of riser height and 34% of step depth inconsistencies were greater than 9.525 mm (Cohen et al., 2009). Francksen et al., (2022) found that adults were able to adjust their stepping behavior for increases in depth, but they could not adjust for step riser height variation over 10 mm (Francksen et al., 2020). This suggests that inconsistent step risers (> 10 mm), could increase one’s risk of a trip in ascent, or over-stepping during descent (Francksen et al., 2020). These findings align with Cohen (2009), who found that stairway fall injury cases involving excessive riser height were 25% more common than those involving excessive depths. In addition, in that investigation, inconsistencies of > 9.525 mm were observed more often in riser height (60%) compared to inconsistent step depth (34%) (Cohen et al., 2009).

We observed that 80% of fall-related events on stairs occurred when riser height varied by 14 mm and depth varied by 38 mm. Since both inconsistent riser height and depth were present in our observational design, we are unable to distinguish which of these factors may have had a greater role. We did find that when both inconsistencies were present, there was a greater frequency of fall-related events.

While older stairways could have greater inter-step inconsistencies (these stairways were built in 1971), the 1967 Uniform Building Code (Sec. 3305(d)) that was in place in the United States states the maximum variations in the height of risers and width of treads in any one flight shall be 3/16 in (4.76 mm). Although in this case, inter-step variations appear to exceed the guidelines that demonstrates a secondary challenge in code reinforcement.

For stairways that had greater inter-step inconsistencies, we assessed whether the addition of high-contrast stripes would reduce the likelihood of fall-related events. Our data suggest that the addition of high-contrast stripes may reduce fall-related events on stairs when large inter-step inconsistencies are present. Only 3 of 16 (19%) fall-related events in this study occurred on a stairway with high-contrast striping compared to 13 of 16 (81%) events on the control stairway without such enhancements.

While the combined high-contrast striping intervention, also known as a horizontal-vertical illusion, appears to have reduced the likelihood of fall-related events on stairways, its mechanism remains unclear. It is possible that the horizontal-vertical illusion alters the precision by which the step’s metrics are estimated, without increasing attention to the stairs. Another possibility is that the novelty of the intervention commands greater attention to the stairs resulting in better estimation of the metrics of the stairs. Indeed, the novelty of the additional contrast, rather than the horizontal-vertical illusion itself, could have drawn attention to the stairs resulting in a reduction of fall-related events. Therefore, it is possible that signage alerting stair users of the inconsistencies may have the same result.

Alternatively, Schomaker et al., (2012) suggests that novel, visual stimuli, such as a change in contrast, may increase attention to the stairs (Weissgerber et al., 2015). It is worth noting that the horizontal-vertical illusion (striping intervention), and the counterbalanced intervention in our investigation were installed several days before each data collection began. It is likely that some of the stair users had exposure to the striping prior to the start of video recording. This early exposure could have reduced the novelty of the striping in those offered the opportunity, thus reducing attention drawing effect. This also true for those who traversed the stairs regularly due to classes in the building. Perhaps this provides additional support for the ‘enhanced precision of our estimate of the steps’ edge effect’ of the horizontal-vertical illusion via high-contrast striping. The high-contrast striping intervention utilized included both black vinyl strips applied to the step’s edges and black-and-white vertical stripes on the last and top steps’ faces. Together, the combined striping intervention with abutting edges, formed ‘T’ configurations which could have been a necessary component to increase perceived step riser height (Schofield, 2022). In further support, the horizontal-vertical illusion effect is reduced when only edge highlighters are present (Skervin et al., 2021). Indeed, our results suggest that horizontal-vertical style illusion is associated with a smaller distribution of fall-related events. It is possible though that other approaches such as the Müller-Lyer illusion could lead to similar outcomes. Recent, outdoor, observation research suggests that greater vertical foot clearance occurs when a ‘fins out’ configuration is applied to a two-step stairway (Shim et al., 2022), and this illusion could address both varying, short and taller, riser height step inconsistencies.

While these results are promising, several limitations of this study should be acknowledged. Since most of the observations in this study were younger adults, future work should consider targeting older or clinical populations (e.g., those with visual impairment) to determine how inter-step inconsistencies impact these populations and whether an intervention similar to what was used here could have a larger impact on fall reduction than in younger adults. In addition, since both inconsistent riser height and tread depth steps were present in our observational design, we are unable to distinguish which had the greater impact. Therefore, future observational designs should assess these two factors independently. In a similar vein, the *combined* high-contrast striping intervention was associated with a smaller distribution of fall-related events. Thus, future observational work may consider assessing variations of the horizontal-vertical illusion to assess whether it is the aggregated effect of the edge highlighter and vertical striping that influences a step illusion effect.

## 5. Conclusions

Overall, it is improbable that the observed distribution of fall-related events occurred by chance and therefore these data provide support that contrast enhancement (i.e., striping) may counteract the increased fall-risk associated with inter-step inconsistencies. While the mechanisms of its action remain unclear, the addition of high-contrast striping appears to have a positive impact on the incidence of fall-related events in the presence of inter-step inconsistencies.

## Data Availability

Data produced in the present study is available upon reasonable request to the authors.

